# COVID-19: a retrospective cohort study with focus on the over-80s and hospital-onset disease

**DOI:** 10.1101/2020.05.11.20097790

**Authors:** Simon Brill, Hannah Jarvis, Ezgi Ozcan, Thomas Burns, Rabia Warraich, Lisa Amani, Amina Jaffer, Stephanie Paget, Anand Sivaramakrishnan, Dean Creer

## Abstract

**Objectives:** To describe the presenting features and outcomes of patients with COVID-19 in a UK hospital, with a focus on those patients over 80 years and patients with hospital onset infection.

**Design:** Retrospective cohort study with data extracted from the electronic records of patients with PCR-confirmed COVID-19 admitted to our institution.

**Setting:** Suburban general hospital serving London’s most populous borough.

**Participants:** The first 450 inpatients admitted to our hospital with swab-confirmed COVID-19 infection.

**Primary outcome:** The primary outcome measure was death during the index hospital admission.

**Results:** The median (IQR) age was 72 (56, 83), with 150 (33%) over 80 years old and 60% male. Presenting clinical and biochemical features were consistent with those reported elsewhere. The ethnic breakdown of patients admitted was similar to that of our underlying local population with no excess of BAME deaths. Inpatient mortality was high at 38%.

Patients over 80 presented earlier in their disease course and were significantly less likely to present with the typical features of cough, breathlessness and fever. Cardiac co-morbidity and markers of cardiac dysfunction were more common, but not those of bacterial infection. Mortality was significantly higher in this group (60% vs 28%, p < 0.001).

31 (7%) of patients were classified as having hospital-onset COVID-19 infection. The peak of hospital-onset infections occurred at the same time as the overall peak of admitted infections. Despite being older and more frail, the outcomes for this cohort were no worse.

**Conclusions:** Inpatient mortality was high, especially among the over-80s, who were more likely to present atypically. The ethnic composition of our caseload was similar to the underlying population. While a significant number of patients presented with COVID-19 while already in hospital, their outcomes were no worse.

**Strengths and Limitations of this Study:**

- This study captures almost 80% of the admitted cases in our institution providing an accurate representation of the experiences of a London hospital during the early peak of the COVID-19 pandemic
- The focus on the clinical and biochemical presentation and outcomes in patients over 80 years of age has a high relevance to UK population which is older and frailer than previously reported cohorts from elsewhere
- The ethnicity of patients admitted to our hospital was similar to that of the underlying local population
- To our knowledge this study is the first to report the prevalence and outcomes of hospital onset disease in the UK
- This study is subject to the usual limitations of retrospective observational research, including a proportion of missing data

## Introduction

In December 2019, a febrile respiratory tract illness was reported in a cluster of patients in Wuhan City (Hubei Province, China)^1^ which we now recognise as a novel pathogenic strain of coronavirus (SARS-coronavirus-2 [SARS-CoV-2])^2^. The World Health Organisation (WHO) subsequently declared coronavirus disease 2019 (Covid-19) a public health emergency of international concern^3^. The infection has spread rapidly across the globe with nearly 3 million infections reported worldwide and nearly 200,000 deaths by 25^th^ April^4^.

The first laboratory-confirmed case of COVID-19 in the United Kingdom was reported on January 30^th^ 2020^5^ with a subsequent rapid rise in the number of cases nationally. As of the 1^st^ May 2020,177,454 patients have tested positive and of those hospitalised in the UK who tested positive for coronavirus, 23,229 have died^6^. Infection numbers peaked in London two to three weeks ahead of much of the rest of the UK.

The first COVID-19 cases in Barnet were reported on 5th March 2020. Barnet is the most populous London Barough with a 2017 estimate of 406,600 inhabitants^7^, and an elderly population with a large number of care homes. Barnet Hospital, a busy suburban hospital with 440 beds, confirmed its first PCR positive COVID-19 patient on 9th March 2020. Since this initial case numbers have risen rapidly with the number of laboratory confirmed COVID-19 inpatients admitted at 587 by 25^th^ April. The peak daily number of positive tests on inpatients was 48, on 2^nd^ April, and the number of confirmed COVID-19 inpatients peaked at 274 on the 6^th^ April.

Since the early case reports there have been numerous publications from China^8^, the US^9^ and elsewhere^10^ describing presenting features and outcomes of the disease and, more recently, from the UK^11^. However, few prior publications have examined the presentation of Covid-19 in the over-80s and none have reported rates of hospital-onset infections. There are also significant concerns in the UK about an apparent excess in COVID-19 related mortality among ethnic minorities^12^.

We therefore report here a detailed profile of our first 450 laboratory-confirmed cases of Covid-19. This comprises 77% of our cases as of 25^th^ April. We examine the demographics, ethnicity, clinical and biochemical features, presentations in older adults and hospital-acquired infections. This will provide useful information as services in the UK are remodeled in the run-up to lifting of restrictions and a possible second peak of infections.

## Methods

Inpatients returning consecutive positive polymerase chain reaction (PCR) tests for SARS-CoV2 on nasopharyngeal, nose or throat swabs during their hospital admission were included for analysis. Data were collected retrospectively from the electronic patient record. Patients with a clinical diagnosis of COVID-19 without PCR confirmation were not included.

Standardised data were collected on demographic features, ethnicity and the presence of co-morbidities (prior diagnosis of cardiac disease [any], hypertension, diabetes, respiratory disease [any], and immunosuppression). In those patients over 65 years of age the Clinical Frailty Score (CFS)^13^ was recorded where available. The presence of care needs prior to admission, including carers at home and institutional care, was recorded.

Community-onset infection was defined as a positive test within 14 days of hospital admission and hospital-onset infection if the patient had continuously been an inpatient for 14 days prior to the positive PCR test. Data were recorded at the point of presentation, defined as the day of hospital admission (community-acquired infections) or documentation of first symptom presentation in the medical notes (hospital-acquired infections). Clinical data included symptom duration, and presenting symptoms and signs. Fever was defined as a temperature > 37·8 Celsius.

Biochemical data included serum lymphocyte and neutrophil counts, C-reactive protein (CRP), procalcitonin, cardiac troponin T, lactate, D-dimer, and glucose. The presence of acute kidney injury was defined according to 2012 Kidney Disease: Improving Global Outcomes guidance^14^. These tests were analysed by the hospital clinical laboratory; details including normal limits and detection thresholds are included in the supplementary appendix (Table S1). Values outside the detection thresholds were entered at the threshold.

The primary outcome assessed was death vs discharge from hospital at the end of the hospital episode, where the patient had reached this point. Some ventilated patients were transferred to other centres including NHS Nightingale and outcome data was unavailable at the time of analysis. Early outcomes at Day 5 following presentation were also captured and defined as discharged, non-intubated inpatient, intubated inpatient, or dead. Other outcomes included length of stay and whether antibiotics were given.

Most variables were not expected to be normally distributed and non-parametric tests were used throughout. Continuous between-group variables were analysed using the Wilcoxon signed-rank test. The Bonferroni correction for multiple analyses was used for comparisons within tables. Categorical variables were analysed using the chi-squared test. Analysis was performed using R Statistics version 3·6·3. Missing data were not imputed; a summary table can be found in the Supplementary Appendix (Table S5).

The data presented here were collected during routine clinical practice and formal Research Ethics Committee review was not required. Approval for the study was granted by the chair of the Trust Clinical Ethics Committee. The full data can be accessed at (to be confirmed).

### Patient and public involvement

This was data routinely collected during clinical practice, and as such patient and public involvement was not required.

## Results

### Demographics, Clinical Characteristics and Outcomes

476 positive swabs were identified; 26 were excluded as they were either too young (less than 16 years old) or were not admitted as inpatients to the hospital. 450 inpatients who underwent consecutive PCR tests confirming Covid-19 between 10^th^ March 2020 and 8^th^ April 2020 were analysed. This represents 77% of the PCR-positive caseload admitted to our hospital to date.

Table 1 displays the demographic and clinical characteristics of the patients, subdivided by outcome. The median (IQR) age was 72 (56, 83) years and, in keeping with the elderly population local to our hospital, 70% (313) were over the age of 60 and 33% (150) over 80. There was a male predominance. Patients who died were significantly older (median (IQR) age 80 (72, 88) vs 61 (49, 79), p < 0·001) and more likely to be receiving care in the community (69 (40%) vs 45 (19%), p < 0·001), with a trend towards greater frailty by CFS. 31 of these patients (7%) developed clinical features of Covid-19 while already admitted to hospital for ≥ 14 days.

**Table 1:** Demographics and baseline characteristics by outcome of hospital admission. Comparisons between those who died and those who were discharged used the Kruskal-Wallis test or the Chi-squared test as appropriate. The Bonferroni method was used to correct for multiple comparisons and therefore a stringent p-value cutoff of 0·05/25 = 0·002 was used to assess significance (indicated by *). ** Differences were assessed by χ^2^ test to examine differences in overall composition between groups.

|  |  | By outcome of hospital stay, where available (n=410) |  |  |
| --- | --- | --- | --- | --- |
|  | All patients (n=450) | Discharged (n=237) | Died (n=173) | P value for comparison |
| **Demographics** |  |  |  |  |
| Age in years, median (IQR) | 72 (56, 83) | 61 (49, 79) | 80 (72, 88) | <0.001* |
| Age breakdown, n (%) |  |  |  |  |
| <40 | 32 (7) | 29 (12) | 2 (1) | - |
| 40-59 | 105 (23) | 81 (34) | 10 (6) | - |
| 60-79 | 163 (36) | 70 (30) | 71 (41) | - |
| >80 | 150 (33) | 57 (24) | 90 (52) | - |
| Male gender, n (%) | 272 (60) | 134 (57) | 111 (64) | 0.146 |
| BMI, median (IQR) | 26 (24, 30) | 27 (24, 30) | 25 (23, 31.5) | 0.214 |
| Receiving care prior to admission, n (%) | 118 (26) | 45 (19) | 69 (40) | <0.001* |
| CFS if >65yrs, median (IQR) | 5 (3, 6) | 4 (3, 5.5) | 5 (3, 6) | 0.014 |
| Ever smoker, n (% of available data) | 76 (31) | 50 (35) | 20 (26) | 0.228 |
| Hospital-onset COVID-19, n (%) | 31 (7) | 20 (8) | 7 (4) | 0.117 |
| Ethnicity, n (%) |  |  |  | 0.072** |
| Asian | 51 (11) | 31 (13) | 13 (8) | - |
| Black | 33 (7) | 19 (8) | 11 (6) | - |
| White | 265 (59) | 127 (54) | 118 (68) | - |
| Other | 77 (17) | 42 (18) | 26 (15) | - |
| Unavailable | 24 (5) | 18 (8) | 5 (3) | - |
| Swab returning positive, n (%) |  |  |  | 0.59** |
| First | 410 (91) | 214 (90) | 158 (91) |  |
| Second | 34 (8) | 19 (8) | 14 (8) |  |
| Third | 6 (1) | 4 (6) | 1 (1) |  |
| **Pre-existing comorbidities, n (%)** |  |  |  |  |
| Hypertension | 195 (43) | 87 (37) | 90 (52) | 0.0029 |
| Cardiac condition | 141 (31) | 55 (23) | 78 (45) | <0.001* |
| Diabetes | 134 (30) | 68 (29) | 53 (31) | 0.589 |
| Respiratory condition | 85 (19) | 47 (20) | 34 (20) | 1.00 |
| Immunosuppression | 42 (9) | 27 (11) | 13 (8) | 0.251 |
| **Disease Characteristics** |  |  |  |  |
| Symptoms at presentation, n (%) |  |  |  |  |
| Cough | 317 (70) | 173 (73) | 113 (65) | 0.228 |
| Breathlessness | 282 (63) | 142 (60) | 113 (65) | 0.204 |
| Diarrhoea | 70 (16) | 38 (16) | 28 (16) | 1.00 |
| Symptom duration at presentation in days, median (IQR) | 5 (2, 8) | 5 (2, 9) | 4 (2, 7) | 0.109 |
| Signs at presentation |  |  |  |  |
| Respiratory rate | 24 (20, 30) | 23 (19, 28) | 26 (22, 33) | <0.001* |
| SaO<sub>2</sub>, median (IQR) | 94 (90, 96) | 95 (92, 96) | 92 (88, 95) | <0.001* |
| (n[%] measured on supplemental O<sub>2</sub>) | 235 (52) | 106 (45) | 106 (61) |  |
| Heart rate, median (IQR) | 90 (80, 103.5) | 90 (80, 103.5) | 90 (78.25, 100) | 0.970 |
| Systolic BP <100 | 38 (8) | 16 (7) | 19 (11) | 0.180 |
| Median temperature | 37.9 (37.2, 38.4) | 38 (37.3, 38.5) | 37.8 (37.2, 38.1) | 0.004 |
| Fever >37.8 | 262 (58) | 143 (60) | 88 (51) | 0.081 |
| Abnormal chest radiograph, n (%) | 370 (82) | 181 (76) | 153 (88) | 0.015 |
| Antibiotics given, n (%) | 332 (74) | 165 (70) | 136 (79) | 0.049 |
| Length of stay in days, median (IQR) | 7 (3, 11) | 7 (4, 12) | 6 (3, 10) | 0.127 |
| Length of stay in days, median (IQR) | 7 (3, 11) | 7 (4, 12) | 6 (3, 10) | 0.127 |
| Status at Day 5, n (%) |  |  |  |  |
| Discharged | 79 (18) | 79 (33) | 0 (0) | - |
| Non-intubated inpatient | 243 (54) | 146 (62) | 77 (45) | - |
| Intubated | 56 (12) | 10 (4) | 26 (15) | - |
| Died | 70 (16) | 0 (0) | 70 (40) | - |

59% overall were white, similar to the 59·7% of the local population who were classified as ethnically white according to 2020 projections from 2011 census data^7^. There was a trend towards a difference in ethnicity, with a higher proportion of white ethnicity among those who died (χ^2^ p = 0.07), but this reflects documented age-related differences in our local population with a higher proportion of our older residents being white^15^.

Hypertension was present in 43% (195) of patients admitted, with cardiac disease in 31% (141), diabetes in 30% (134), respiratory conditions in only 19% (85) and immunosuppression in 9% (42). Pre-existing cardiac disease (78 (45%) vs 55 (23%, χ^2^ p =0.005) and hypertension (87 (37%) vs 90 (52%, (χ^2^ p = 0.003), but not diabetes, respiratory conditions or immunosuppression were more prevalent in those patients who died than those who were discharged. Further detail may be found in the Supplementary Appendix (Tables S2, S3 and S4).

Cough and breathlessness were reported at presentation in 70% (317) and 63% (282) respectively. Diarrhoea was reported by 16% (70) patients. The median duration of symptoms was 5 (2, 8) days. At presentation, those who died had a higher respiratory rate (26 (22, 33) vs 23 (19, 28), p < 0·001) and lower oxygen saturations (92 (88, 95) vs 95 (92, 96), p < 0·001) than those discharged. Presenting temperature, heart rate and the presence of hypotension were no different between these groups although there was a trend towards lower median temperature in those who died. The majority (74%) of patients received antibiotic therapy for a median (IQR) duration of 5 (3, 7) days.

At Day 5 following admission 18% (79) of patients had been discharged, 54% (243) were not intubated but remained in hospital, 12% (56) were intubated and receiving mechanical ventilation and 16% (70) had died. Of those intubated at Day 5, 46% (26) had died, 18% (10) gone home, 13% (7) remained in hospital and 23% (13) had unknown outcomes, usually following transfer to another centre (Figure 1). The median length of stay was 5 (2, 8) days.

**Figure 1:**
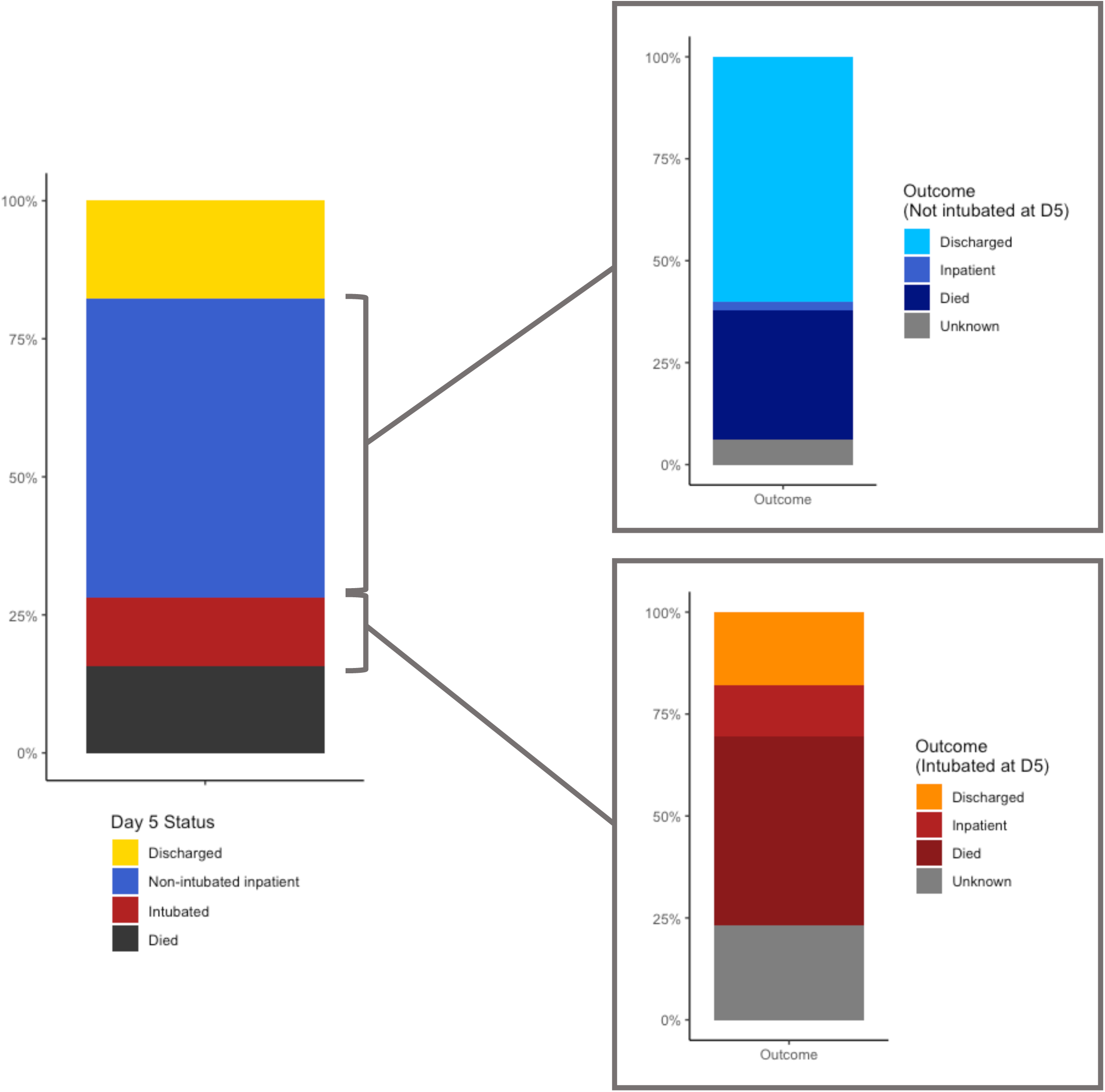
Outcomes for first hospital admission by status at Day 5.

At the time of data analysis 38% (173) patients had died, 53% (237) had been discharged, 3% (12) remained in hospital and 6% (28) had unknown outcomes.

### Biochemical disease characteristics by outcome

Table 2 summarises the biochemical presenting features overall and by outcome of hospital stay.

**Table 2.** Laboratory studies at presentation with Covid-19, subdivided by the outcome of the hospital stay where available (n=410). Comparisons between those who died and those who were discharged used the Kruskal-Wallis test or the Chi-squared test as appropriate. The Bonferroni method was used to correct for multiple comparisons and therefore a stringent p-value cutoff of 0·05/12 = 0·0042 was used to assess significance (indicated by *).

|  |  | **By outcome of hospital stay,**<br>**where available (n=410)** |  |  |
| --- | --- | --- | --- | --- |
|  | **All patients**<br>**(n=450)** | **Discharged**<br>**(n=237)** | **Died**<br>**(n=173)** | **P value for**<br>**comparison** |
| Lymphocyte count | 0.84 (0.58, 1.23) | 0.91 (0.67, 1.28) | 0.78 (0.535, 1.14) | 0.009 |
| Neutrophil count | 5.72 (3.84, 8.61) | 5.32 (3.48, 7.82) | 6.6 (4.178, 9.750) | 0.001* |
| Neutrophil:<br>lymphocyte ratio | 221.5 (112.8, 333.2) | 217 (117, 323) | 239 (106.5, 343.5) | 0.676 |
| CRP | 99 (46, 176.5) | 68 (31, 140) | 131 (74, 199) | <0.001* |
| CRP > 100, n (%) | 221 (49) | 83 (35) | 108 (62) | <0.001* |
| Procalcitonin | 0.26 (0.13, 0.73) | 0.20 (0.11, 0.44) | 0.37 (0.17, 1.35) | <0.001* |
| Troponin | 23 (9, 50) | 12 (6, 33) | 42 (20, 71.5) | <0.001* |
| Lactate | 1.3 (0.9, 1.8) | 1.2 (0.9, 1.6) | 1.5 (1.1, 2.25) | <0.001* |
| D-dimer | 1294 (616.5, 2429.8) | 1186 (535, 2340) | 1577 (814, 2548) | 0.014 |
| Glucose | 6.6 (5.8, 8.3) | 6.5 (5.575, 8.0) | 6.9 (5.9, 8.625) | 0.017 |
| Acute kidney injury,<br>n(%) | 85 (19) | 24 (10) | 54 (31) | <0.001* |

Inflammatory markers that differed significantly between those that died and those that were discharged were neutrophil count (6·6 (4·178, 9·750) vs 5·32 (3·48, 7·82), p = 0·001), CRP (131 (74, 199) vs 68 (31, 140), p < 0·001) and procalcitonin (0·37 (0·17, 1·35) vs 0-20 (0·11, 0·44), p <0·001. Cardiac troponin (1422 (506, 4473) vs 12 (6, 33), p < 0·001) and lactate (1·5 (1·1, 2·25) vs 1·2 (0·9,1·6), p <0·001). Acute kidney injury (10% (24) vs 31% (54), χ^2^ p < 0·001)) was more prevalent in those who died.

### COVID-19 in the over-80s

33% (150) of patients were aged 80 years or over and the characteristics of COVID-19 in these patients was examined. The median (IQR) age in this group was 86 (83, 91) and the oldest was 101. 53% (70) were male.

When compared with those patients under 80 years, these patients were more frail (median (IQR) CFS 5 (4,6 vs 3 (2, 5), p <0·001)), more likely to have been receiving care prior to admission (51% (76) vs 14% (42), χ^2^ p <0·001), and had lower BMI (median (IQR) 24 (21, 40) vs 28 (25, 32), p <0·001). Ethnicity breakdown was significantly different in the over-80s, (χ^2^ p <0·001)), with the proportions of White patients (72%) higher than in the overall caseload, similar to the age-related ethnic composition of our older local population.

Prior diagnosis of a cardiac condition was significantly more common in those >80 years (53% (79) vs 21% (62), χ^2^ p <0·001), as was hypertension (51% (76) vs 40% (119), χ^2^ p = 0·0) but diabetes, respiratory conditions and immunosuppression were not.

Patients over 80 were significantly less likely to present with the typical syndrome of breathlessness (48% (72) vs 70% (210), χ^2^ p <0·001), cough (58% (87) vs 77% (230), χ^2^ p <0·001), or fever (50% (75) vs 62% (186), χ^2^ p = 0·02). Median (IQR) respiratory rate was lower at presentation (23 (19, 28) vs 26 (20, 32) p = 0·001), as was heart rate (84·5 (77·25, 96·75) vs 95 (80,106), p < 0·001). The median (IQR) symptom duration prior to presentation was significantly lower in the older group (4 (1, 6·25) vs 7 (2, 9) days, p <0·001).

Biomarker profiles were compared between the older and younger groups for those biomarkers listed in Table 2. Median (IQR) troponin (56 (35·5, 94·5) vs 14 (7, 32), p < 0·001), D-dimer (1966 (1136, 2768) vs 1120 (538, 2058), p < 0·001), and lactate (1·45 (1·1, 2·1) vs 1·3 (0·9, 1·7) were significantly higher in the over-80s while the lymphocyte count was lower (0·76 (0·53,1·14) vs 0·89 (0·62 vs 1·28, p = 0·007). Notably there were no differences in CRP, procalcitonin, neutrophil count, or glucose between age groups.

Mortality was significantly higher in the older age group (60% (90) vs 28% (83), χ^2^ p <0·001). However, of the over-80s, those that died were not significantly older than those that did not (median (IQR) age 87·5 (83·25, 91) vs 86 (82·75, 90·25), p = ns), although they were more frail (median (IQR) CFS 6 (5, 7) vs 5 (4, 6), p = 0·002). Median (IQR) respiratory rate (24 (20, 30) vs 21 (18, 25.25), p < 0·001) and heart rate (88 (78, 99) vs 83 (74, 88), p = 0·03) were significantly higher in those that died. Median (IQR) CRP was significantly higher in those that died than those that survived (125 (73·25,199·75) vs 67·50 (25·75,119), p < 0·001)); other biomarkers were not significantly different.

### Hospital-acquired infections

7% (31) of infections were hospital-onset. The median (IQR) duration of hospital stay prior to Covid-19 testing was 20 (14, 36) days. The first hospital-onset infection was recorded 8 days after the first positive test on an inpatient; the peak of hospital-onset infections occurred approximately 3 weeks later and mirrored that of the community-onset infections (Figure 2).

**Figure 2:**
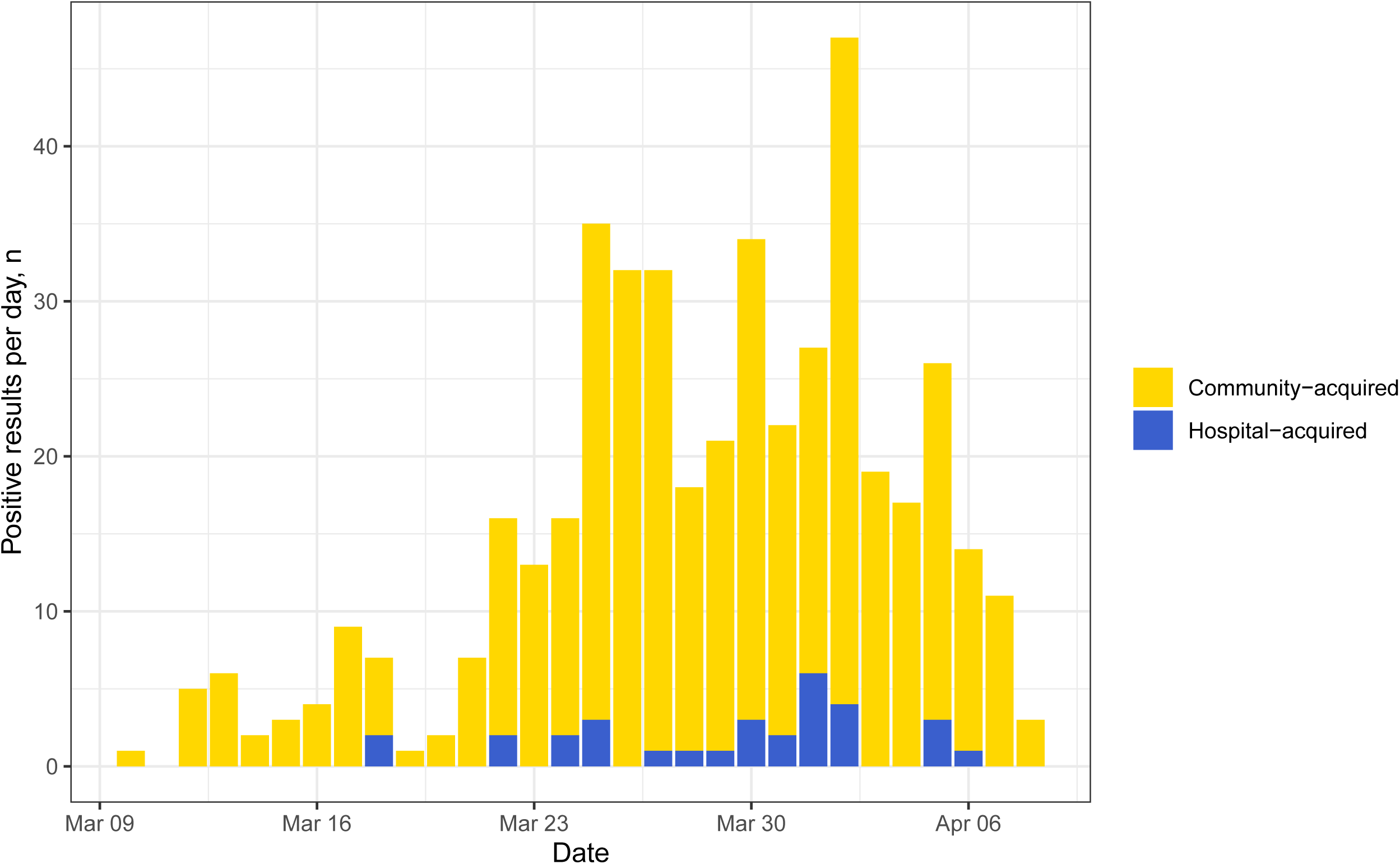
Timeline of cases of community-onset and hospital-onset COVID-19 in patients admitted to hospital.

These patients were older (median (IQR) age 80 (72·5, 88·5) vs 71 (55·5, 83), p = 0·002), slightly more frail (median (IQR) CFS 5 (4, 6) vs 5 (3, 6), p = 0·047) and more likely to have required care prior to admission (55% (17) vs 24% (101), χ^2^ p <0·001). Median (IQR) symptom duration was much shorter than in community-onset infections (1 (1, 2) vs 5 (2, 8) days, p < 0·001) likely reflecting the enhanced monitoring of these hospital inpatients. Median CRP (38·5 (12.25, 72·50) vs 104 (50,127), p < 0·001) was significantly lower in the hospital-onset group.

7 (23%) patients with hospital-onset infections died compared to 40% (166) of those with community-onset infections (χ^2^ p = 0·09) suggesting that despite their vulnerability their overall outcomes were no worse. 32% (10) patients with hospital-onset infections were asymptomatic at the time of their swab, which was performed based low oxygen saturations or pyrexia.

## Discussion

We report here the characteristics of nearly 80% of the patients with confirmed COVID-19 presenting to our busy suburban London hospital during the first six weeks of the UK peak.

The presenting features of COVID-19 in our population were similar to those seen in reports elsewhere in terms of presenting characteristics; cough (70%), fever (58%) and breathlessness (63%). Markers of inflammation at presentation (CRP, neutrophil count, procalcitonin and lactate) were elevated in patients who died compared to those who were discharged from hospital, as was the presence of elevated troponin and acute kidney injury. CRP, in particular, seemed to be markedly associated with death independent of age, and a cutoff of 100 was almost twice as prevalent (62% vs 35%) in those who died. This requires further validation. A high mortality rate of 38% was seen for all inpatients, similar to recently published UK data^11^, with a mortality of 60% in the over-80s.

The ethnic profile of the admitted patients (59% White, 41% Black and minority ethnic [BAME]) was almost identical to that of our underlying population, which in 2020 was projected to be 59·7% White and 40·3% BAME^7^. The percentage of White patients was higher in those who died and was 72% in those over 80 years old, again consistent with the higher proportion of White people in those who are older in the borough^15^. This is hugely topical presently; we did not observe the excess of morbidity and mortality recently reported^12^ among BAME patients.

Our cohort included older, frailer patients than has been seen in other studies^16^. Those patients over 80 were less likely to present with typical symptoms of COVID-19 infection and were more likely to have coexistent cardiac disease and elevated biochemical markers of cardiac dysfunction. 31 (7%) of patients were classified as having hospital-acquired COVID-19 infection, but despite being an older, frailer cohort, outcomes were no worse than those seen in community acquired disease.

The size of this cohort, in addition to the breadth of clinical and biochemical data collected, has allowed us to present a reasonably complete picture of COVID-19 as it presented to our UK hospital. Whilst other studies, notably the ISARIC in the UK^11^, have reported on larger cohorts, there has been a paucity of matched clinical and biochemical data specific to a UK population. In addition, we provide insight into two under-reported groups; the over-80s and those with hospital acquired COVID-19. Despite these strengths, there are several limitations to this study. We did not collect data on some presenting features that are important, notably atypical symptoms at presentation and more detail on pre-existing comorbidities and medications. Furthermore, this was an observational study and therefore data collection was not standardised. Owing to this, and the fact that we only introduced a clinical care bundle specifying laboratory tests two to three weeks into the outbreak, there is a proportion of missing data in some of the biochemical variables. This will have affected our ability to detect more subtle signals, although does not diminish the significance of those we have reported. We also did not have follow-up data on those patients discharged and were therefore unable to assess subsequent deaths or readmissions.

Whilst many of the findings from our study correlate with other UK reports^11^, notably a high inpatient mortality and an increasing risk of death with increasing age and cardiac comorbidity, striking differences were observed compared to non UK studies. This reflects the severity of disease in those hospitalised with COVID-19 in the UK as well as the underlying age of our population which was older with an associated increased frailty. The UK experience therefore differs dramatically from the initial reports from China^16^, with a reported in-hospital mortality of 1·4%. It is also higher than the 21% reported in the USA by Richardson and colleagues^9^, although that population was younger and their follow-up duration shorter meaning that fewer patients may have reached this endpoint by the time of analysis. It seems that, at least in the early stages of the epidemic in Wuhan, all patients with COVID-19 were hospitalised regardless of disease severity. Hospital practice in the UK has been to only admit those patients medically requiring hospitalisation and this is therefore a much sicker cohort overall.

Previous studies of COVID-19 in the older population are small and have used differing variations for ‘elderly’ ranging from 60-65 years^17,18^; even these have included only small numbers of patients that would be classically considered ‘elderly’ in the UK, usually over 80 years of age. Mortality is linked to increasing age and, in line with non-covid disease in the elderly, there has been suggestion^19^ but little evidence that these patients are more likely to present atypically. Our findings confirm this and also demonstrate these patients present earlier in their disease course suggesting lower physiological reserve. Lymphocyte count was lower in the over-80s, possibly reflecting age-related immune dysfunction or more severe disease. While age is independently linked to mortality^11^, above the threshold of 80 years the exact age did not differ between outcomes. Although troponin differed significantly between age groups, consistent with the higher incidence of cardiac disease in the over-80s, it did not differ significantly by outcome in the elderly population. This may suggest that although cardiac dysfunction is present in the older population it is not the cause of death per se, although the smaller numbers here may mean that a statistically significant association may have been missed. CRP was associated with poor outcome in this age group but neither this nor procalcitonin differed between the age categories, suggesting that bacterial infection is not a greater driver of disease in older patients.

The incidence of nosocomial COVID-19 has been estimated in other cohorts^20^ as 44%, far higher than we found in our patients. Data have been of poor quality, however, and information from outside Hubei is lacking. For our analysis we used a stringent definition, including only those patients with continuous inpatient admission for the whole of the 14-day incubation period prior to symptoms, and the true incidence may therefore be higher. These patients had already had prolonged admissions for unrelated reasons and correspondingly were an older and more frail group than the population as a whole. Despite this, CRP was lower and their outcomes were no worse, reflecting the fact that they effectively acquired COVID-19 incidentally while in hospital rather than presenting due to severe infection. The peak incidence of nosocomial infections mirrored the overall peak of community infections and the mode of transmission remains unclear; this may include relatives visiting before they were excluded and asymptomatic infected healthcare workers. It may also have included transmission from other patients either directly or via healthcare workers.

The data presented here contribute to the understanding of COVID-19 as it applies to a UK population. In particular, the focus on differences in the presentation and outcomes in the elderly is of utmost importance given the significantly higher mortality seen in this population and the emerging picture of how COVID-19 has affected care homes. Further research to elucidate the mechanisms by which age related biological variance impacts on the pathogenic response in COVID-19 is vital to enabling effective therapeutic interventions. Likewise, exploring the mechanisms by which COVID-19 infection is transmitted to and acquired by vulnerable inpatients is crucial to enabling robust infection control policy in the ongoing management of this and future pandemics.

## Data Availability

Data will be uploaded; location TBC

## Author Statement

SB, HJ, EO, TB, RW, LA, SP, AS and DC conceived and initiated the project. EO, TB, RW, LA and LA-T extracted the data from electronic case records and performed preliminary data analysis and local presentation. SB performed the final data analysis. SB, HJ, AJ and DC wrote the first draft of the manuscript. All authors contributed to interpreting the data and writing the final paper.

## Acknowledgements

We thank Debbie Bertfield for her helpful input regarding frailty and COVID-19 in the over80s.

## References

1. Huang C, Wang Y, Li X, et al. Clinical features of patients infected with 2019 novel coronavirus in Wuhan, China. Lancet 2020;395:497–506.

2. Lu R, Zhao X, Li J, et al. Genomic characterisation and epidemiology of 2019 novel coronavirus: implications for virus origins and receptor binding. Lancet 2020;395:565–574.

3. World Health Organisation. Coronavirus disease (COVID-19) Pandemic. Available: https://www.who.int/emergencies/diseases/novel-coronavirus-2019. Last accessed 30th April 2020.

4. World Health Organisation. Coronavirus (COVID-19). Available: https://covidl9.who.int/. Last accessed 30th April 2020.

5. UK Government (2020). Coronavirus (COVID-19) in the UK. Available: https://coronavirus.data.gov.uk/?_ga=2.137891512.944000509.1587819030-1391566006.1580594784. Last accessed 30th April 2020.

6. UK Government. Guidance: Number of coronavirus (COVID-19) cases and risk in the UK. Available: https://www.gov.uk/guidance/coronavirus-covid-19-information-for-the-public. Last accessed 2nd May 2020.

7. London Borough of Barnet (2020). Demography. Available: https://jsna.barnet.gov.uk/1-demography. Last accessed 30th April 2020

8. Zunyou Wu, Jennifer M. McGoogan. (2020). Characteristics of and Important Lessons From the Coronavirus Disease 2019 (COVID-19) Outbreak in China: Summary of a Report of 72,314 Cases From the Chinese Center for Disease Control and Prevention. JAMA 2020; 323 (13): 1239–1242

9. Richardson S, Hirsch JS, Narasimhan M, et al. Presenting Characteristics, Comorbidities, and Outcomes Among 5700 Patients Hospitalized With COVID-19 in the New York City Area. JAMA 2020: doi:10.1001/jama.2020.6775

10. Epidemiology for public health. Characteristics of COVID-19 patients dying in Italy, https://www.epicentro.iss.it/en/coronavirus/sars-cov-2-analysis-of-deaths. Last accessed 30th April 2020.

11. Docherty A, Harrison E, Green C, et al. Features of 16,749 hospitalised UK patients with COVID-19 using the ISARIC WHO Clinical Characterisation Protocol. 2020; preprint available at https://www.medrxiv.org/content/10.1101/2020.04.23.20076042v1. Last accessed 2nd May 2020.

12. White C, and Nafilyan V. Coronavirus (COVID-19) related deaths by ethnic group, England and Wales: 2 March 2020 to 10 April 2020. ONS 2020; available at https://www.ons.gov.uk/peoplepopulationandcommunity/birthsdeathsandmarriages/deaths/articles/coronavirusrelateddeathsbyethnicgroupenglandandwales/2march2020to10april2020. Last accessed 8th May 2020.

13. Rockwood K, Song X, MacKnight C, et al. A global clinical measure of fitness and frailty in elderly people CMAJ. 2005 Aug 30; 173(5): 489–495.

14. Kidney Disease: Improving global outcomes. KDIGO Clinical Practice Guideline for Acute Kidney Injury. https://kdigo.org/wp-content/uploads/2016/10/KDIGO-2012-AKI-Guideline-English.pdf. Last accessed 1st May 2020.

15. London Borough of Barnet (2015). Barnet Joint Strategic Needs Assessment, 2015–2020. Available: https://www.barnet.gov.uk/sites/default/files/assets/jsna/Downloads/BarnetsJSNA20152020.pdf. Last accessed 27th April 2020.

16. Guan W, Ni Z, Hu Y et al. Clinical Characteristics of Coronavirus Disease 2019 in China. NEJM 2020; 382: 1708–1720.

17. Liu K, Chen Y, Lin R, Han K (2020). Clinical features of COVID-19 in elderly patients: A comparison with young and middle-aged patients. J Infection (in press).

18. Niu S, Tian S, Lou J, et al. Clinical characteristics of older patients infected with COVID-19: A descriptive study. Arch Gerontology Geriatrics 2020: https://doi.Org/10.1016/j.archger.2020.104058

19. Holroyd-Ledu, J, Gandell D, Miller A, Petrov D. COVID-19 in Older Adults. University of Toronto 2020: https://www.rgptoronto.ca/wp-content/uploads/2020/04/COVID-19-Presentations-in-Frail-Older-Adults-U-of-C-and-U-fo-T.pdf/. Last Accessed 2nd May 2020.

20. Zhou Q, Gao Y, Wang X et al. Nosocomial Infections Among Patients with COVID-19, SARS and MERS: A Rapid Review and Meta-Analysis. 2020; Preprint available at https://www.medrxiv.org/content/10.1101/2020.04.14.20065730v1. Last accessed 2nd May 2020.

